# Congenital transmission of Chagas disease: The role of newborn therapy on the disease’s dynamics

**DOI:** 10.1101/2024.02.18.24302989

**Authors:** Meriem Boukaabar, Bismark Oduro, Paul Chataa

## Abstract

Chagas disease, also known as American trypanosomiasis, is caused by a protozoan blood-borne pathogen called Trypanosoma cruzi. The World Health Organization (WHO) has classified Chagas as one of 21 neglected tropical diseases present in the world and estimates that 6-7 million people are currently infected with Chagas. Congenital transmission of Chagas disease contributes to a significant amount of new infections, especially in endemic areas where 22.5% of new infections are due to congenital transmission. In this paper, we investigate the impact of congenital transmission on the dynamics of Chagas disease through a mathematical model. Specifically, we examine how treatment and the efficacy of the therapy for newborns impact the progression and spread of Chagas disease.

The influence of newborn therapy on the dynamics of the model is thoroughly investigated, both theoretically and numerically. The results illustrate the importance of an effective treatment for newborns in reducing infected cases of the Chagas. We observed that if vector transmission can be controlled, then at least 41% of the newborns need to be treated to curb the disease, and varying the newborn treatment rate and its efficacy significantly shapes the disease’s spread. The finding further shows that the therapy given to newborns is not sufficient but necessary to curb the transmission of Chagas disease, and a comprehensive approach that includes vector and vertical transmission control strategy is essential for eradicating Chagas disease.

## Introduction

Chagas disease, also known as American trypanosomiasis, is an anthropozoonosis disease caused by a protozoan blood-borne pathogen called Trypanosoma cruzi [14]. The disease is predominantly active in Latin America, where it is a major public health issue [3]. The World Health Organization (WHO) has classified Chagas as one of 21 neglected tropical diseases in the world [8]. Additionally, the WHO estimates that 6-7 million people are currently infected with Chagas, and 75 million people are at risk for acquiring the disease [10]. Higher incidence rates are typically associated with areas that have poorly constructed housing, which serve as hiding places for the insect vectors that transmit the disease [3]. The disease is vectorized by Triatomine (reduviid) bugs, also known as “kissing bugs,” because they bite the host around their lips when they feed [17]. When the bug feeds on humans, it defecates, which allows the T. cruzi to exit with the feces and enter the host’s body [13]. This is one of the most common routes of infection. Other common routes of infection include congenital transmission, consumption of triatomine insects, needle sharing, and transfusional transmission [19]. The highest number of new acute infection cases comes from vector and congenital transmission [9]. This is especially true in Latin American countries, where approximately 22.5% of new infections are due to congenital transmission [6]. In both the acute and chronic phases of the infection, the disease can be transmitted from the infected mother through the placenta to the embryo or fetus [11]. In 1 *−* 10% of infants of infected mothers, congenital T. cruzi infection occurs [3]. Pregnant-infected women typically have higher rates of premature births and miscarriages [7]. Typically, the mothers and the infected children are asymptomatic, which makes the diagnosis of Chagas challenging [5]. Even in cases where symptoms are present, they are non-specific symptoms like fevers, swollen lymph nodes, and hepatosplenomegaly [21]. Regardless of the symptoms, all untreated infected infants are at a 20 *−* 30% risk of developing severe cardiac and intestinal complications later on in their life [2]. Ultimately, the faster the diagnosis and subsequent treatment, the more effective it is [11]. There is a 90 *−* 95% cure rate when the disease is recognized early, and treatment is used [20]. As infected patients age, the cure rate decreases, so diagnosis should be a priority [16]. Current diagnosis and treatment methods consist of a multistep method. Firstly, detection requires maternal serological screening [18]. Typically, if a mother tests positive two times in a row, the newborns are suspected of having Chagas disease and are tested [3]. The most common method for testing newborns is examining cord blood from their seropositive mothers using microscopy (also called the ”micro method”) or polymerase chain reaction (PCR) techniques, anytime until they are one-month old [15]. Infants who test positive via microscopy or PCR are considered to have Chagas disease [18]. Unfortunately, these methods are unreliable; more than 50% of infections are not recognized by microscopy [12]. Therefore, infants who are tested after one month of birth or test negative are retested using serology when they are 9-12 months old [3]. Treatment options include Benznidazole or Nifurtimox [10]. Treatment during pregnancy is not currently recommended because of a lack of data on safety [16]. However, since the treatment options are highly effective for newborns, treatment should begin as soon as the diagnosis is made [3]. Over the past couple of decades, there have been stricter control measures and detection policies for Chagas disease [1]. Despite these measures, incidence rates have increased in non-endemic regions like Europe and North America due to migration from rural to urban areas [1]. The disease is no longer confined to Latin America and is now a worldwide issue [4]. As such, increased diagnosis and better treatment of the disease is only a small step forward in eradicating congenital Chagas disease. Significant government and health policies must also be put in place to curb this disease. Furthermore, improving education and awareness regarding Chagas disease is crucial for healthcare workers. Combining these strategies will hopefully minimize the prevalence of Chagas disease globally.

A few articles have been published on using mathematical models to explore Chagas disease dynamics, including [23–25, 28]. Raimundo et al. in [26] focused on the congenital transmission of Chagas disease in populations where vectorial transmission has been eliminated. By considering both vertical transmission and the presence of vectorial transmission, our study expanded on this work. Furthermore, we also incorporated vector control measures into the model. Coffield et al. in [27] used a mathematical model to explore Chagas disease transmission by including congenital and oral transmission modes in humans and domestic mammals. The research concluded that while congenital transmission has a limited impact on infection, oral transmission in domestic mammals significantly contributes to the disease’s spread, highlighting the importance of considering alternative transmission modes in disease control strategies [27]. None of these published papers explored the impact of congenital transmission and newborn therapy in controlling the disease; this emphasizes the significance of our study in filling this research gap.

## Materials and methods

In this section, we develop a compartmental model that reflects the dynamics of Chagas disease in both human and vector populations. The vector population is divided into two classes at time *t*: susceptible vectors (*S*_*υ*_) and infected vectors (*I*_*υ*_). The human population is divided into four classes at time *t*: infected acute humans (*I*_*a*_), infected chronic humans (*I*_*c*_), susceptible humans (*S*_*h*_), and newborn babies from infected mothers (*M*). The natural death rate of vectors is denoted by *μ*_*υ*_, and the model assumes all newborns from non-infected mothers are susceptible. If a susceptible vector feeds on an infected acute or infected chronic human, the rate of disease transmission from the human to the vector is represented by *β*_*hυ*_, and the susceptible vector moves to the infected vector class and stays there for life. When an infected vector bites a susceptible human, the disease is transmitted at a rate of *β*_*υh*_, and the susceptible human is moved to the infected acute stage. From there, the infected acute human can progress to the infected chronic human class; this progression rate is denoted by *k*. Individuals in the infected chronic class remain in that class for life unless they leave the population through natural death at rate *μ*_*h*_ or the death rate from Chagas given by *δ*_*h*_. The birth rate of infected acute and infected chronic mothers combined, represented by *b*_*h*_(*I*_*c*_ + *I*_*a*_), is what comprises *M* class. If properly and fully treated, newborns from infected mothers can be moved to the susceptible human class at the rate of *α* = *ωr* where *r* is the treatment rate newborn, and *ω* is the treatment efficacy. Newborns who are not properly treated become classified as infected acute humans at (1 *− α*). A diagram depicting the dynamics explained in this section is shown in Figure 1.

**Fig 1.**
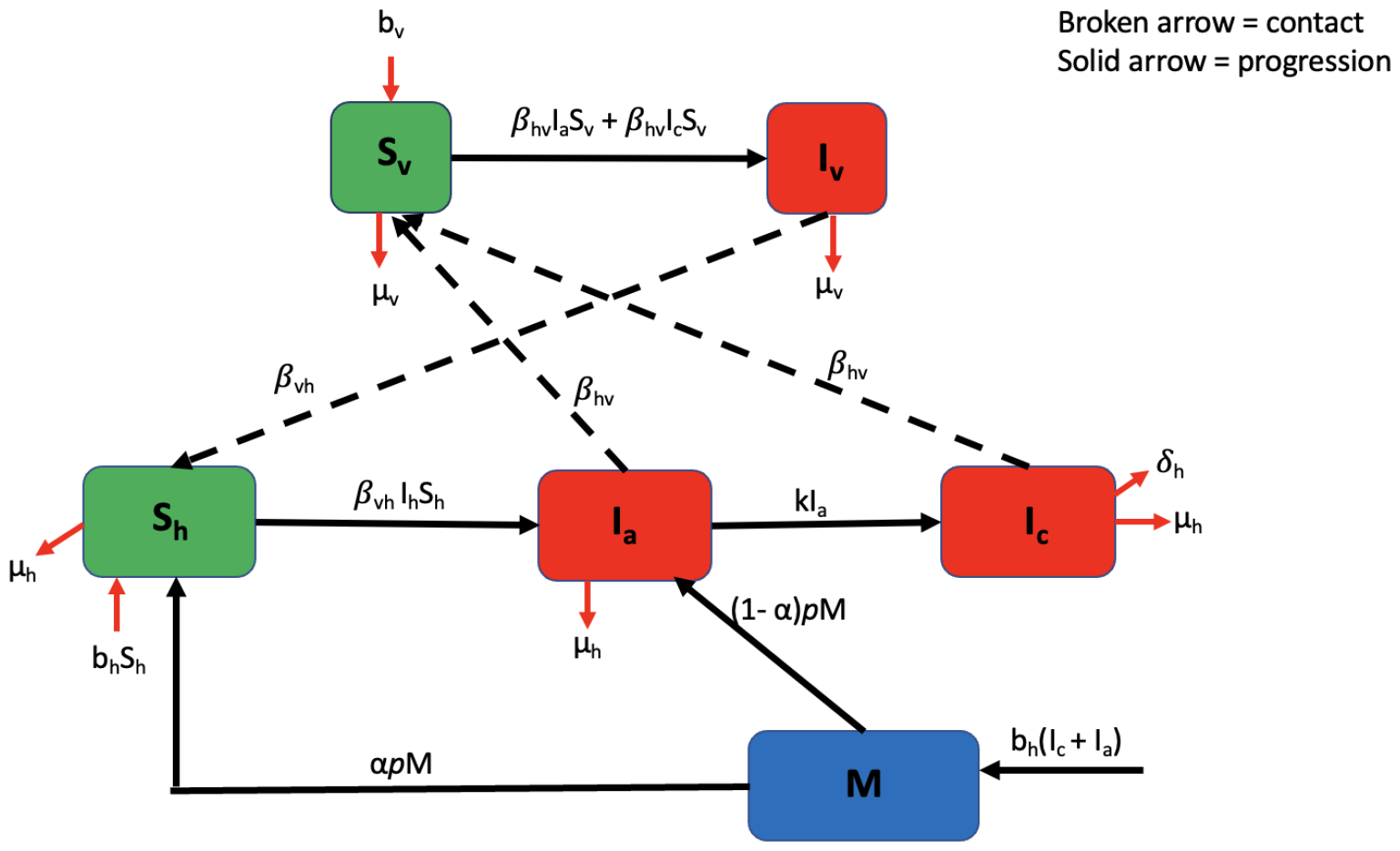
Schematic diagram of the model. See Table 1 for meanings of parameters/variables.

Under the assumptions described above and the diagram, we obtain the following system of nonlinear ordinary differential equations.

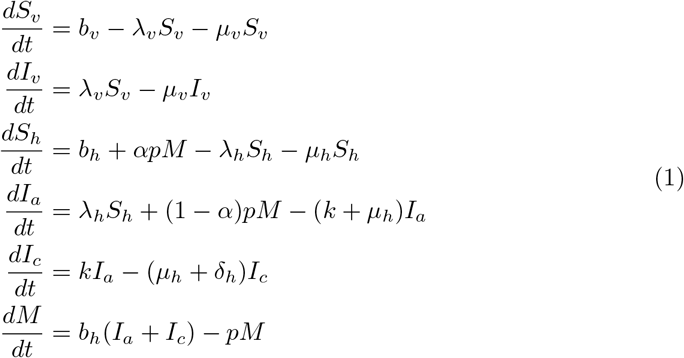

Where

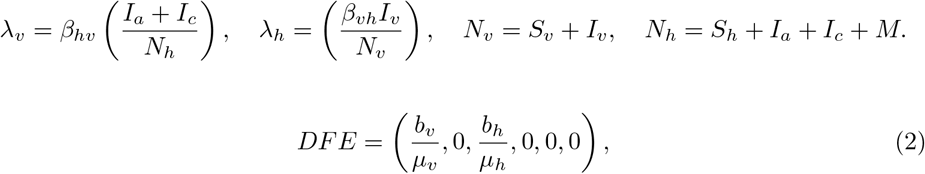

**Table 1.**
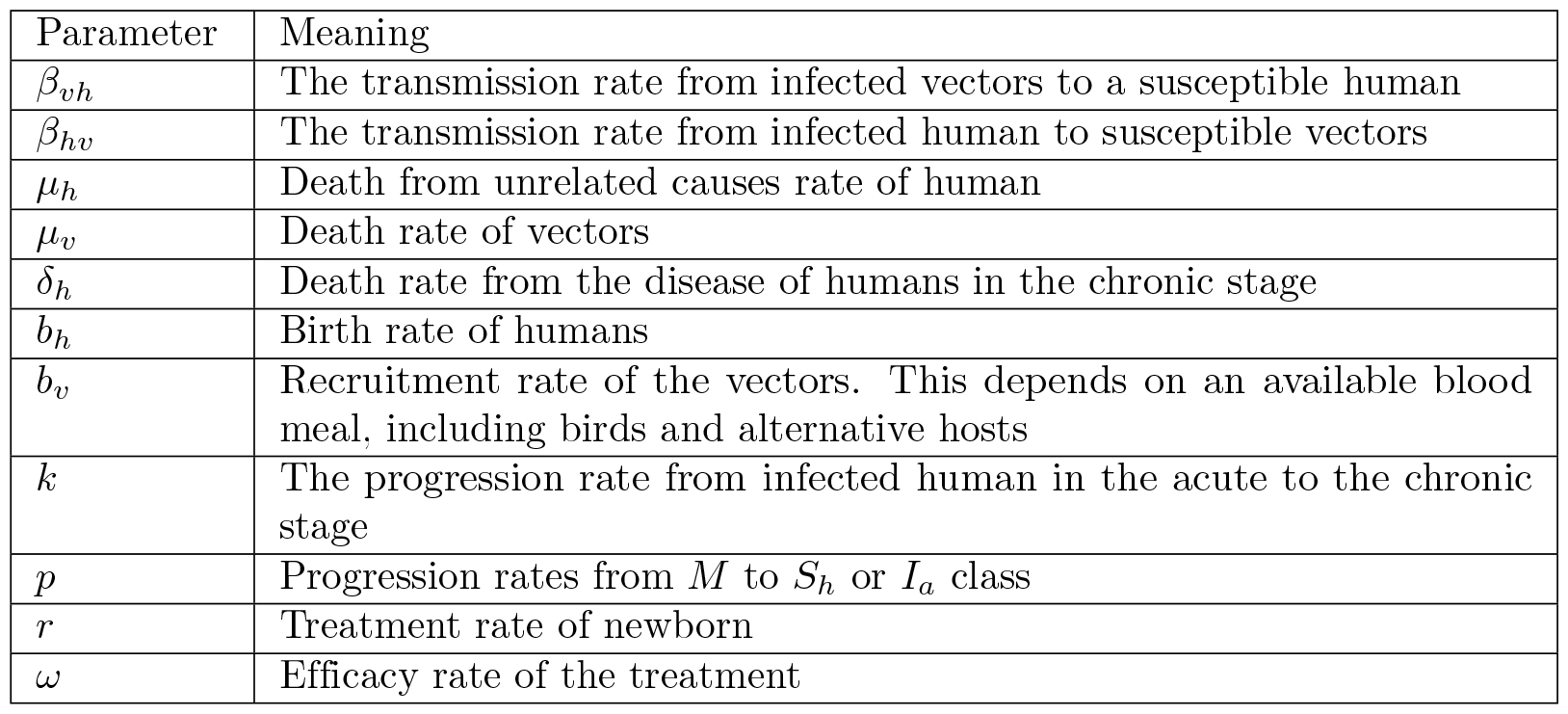
Parameters of the disease model and their meanings.

A next-generation approach is defined as the dominant eigenvalue (spectral radius) of the matrix *FV* ^*−*1^ [30–32], where *F* and *V* ^*−*1^ are matrices determined as:

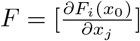 and 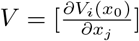. Here, *x*_*j*_ is the number of infested units, *x*_0_ is the disease-free equilibrium, *F*_*i*_ is the rate of appearance of new infection in the infected compartments, 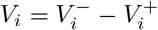 with 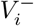 denoting the rate at which infected individuals are transferred out of the infected compartments and 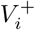 denoting the rate at which individuals are transferred into the infected compartments.

We will use the next generation method to compute the control reproduction number

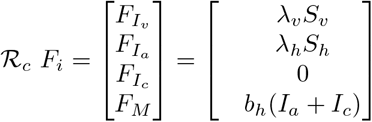

and

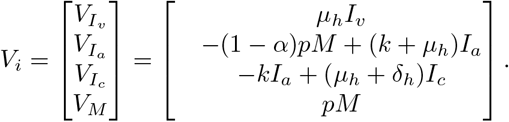

Therefore

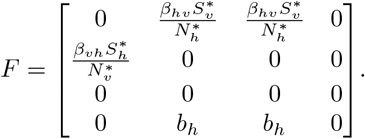

and

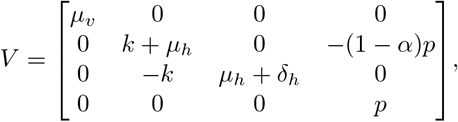

where 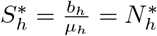 and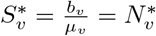.

By the next generation method, the reproduction number is the spectral radius *FV* ^*−*1^. That is,

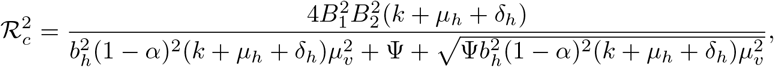

where

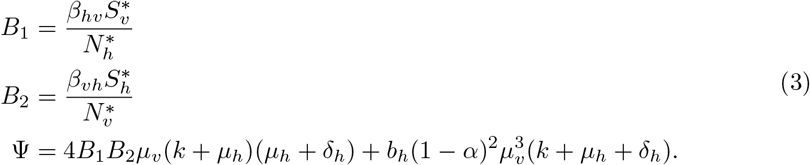

Let us consider a scenario that allows a perfect treatment that is *α* = 1.

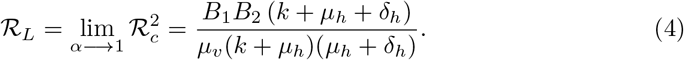

Equation 4 shows that perfect treatment of newborns is insufficient to exterminate the infection if *ℛ*_*L*_ *>* 1. In addition to the newborn therapy, control measures that would reduce *ℛ*_*L*_ below unity are required to eradicate the disease.

## Stability Analysis Results

In this section, we determine the local and global stability of the disease-free equilibrium.

First, we determine the local stability of the disease-free equilibrium by computing the eigenvalues of the linearized Jacobian matrix at the disease-free equilibrium and obtain

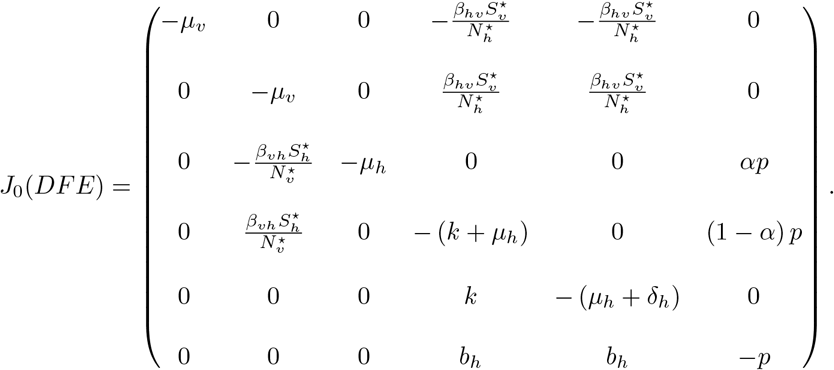

From the Jacobian matrix *J*_0_(*DFE*), the first two eigenvalues are obtain to be *ϱ*_1_ = *−μ*_*v*_ *<* 0, *ϱ*_2_ = *−μ*_*h*_ *<* 0. The remaining four eigenvalues are given by the 4 *×* 4 matrix

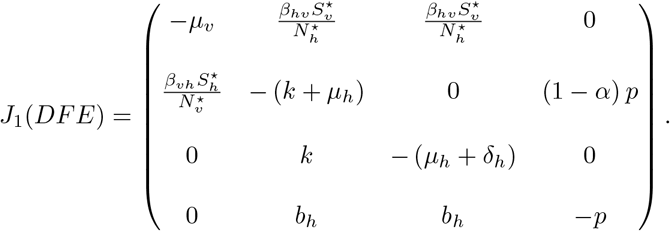

Consider 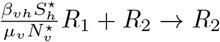, we have

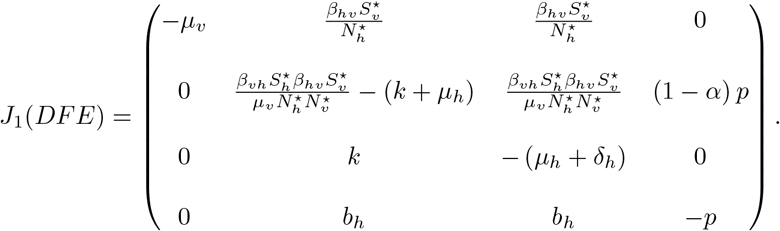

It follows *J*_1_(*DFE*) that

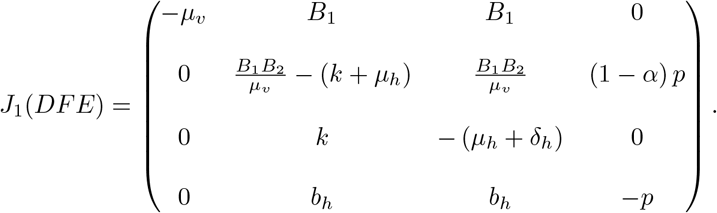

From column 1, the eigenvalue is *ϱ*_3_ = *−μ*_*v*_ *<* 0. The remaining three eigenvalues are given by the 3 *×* 3 matrix

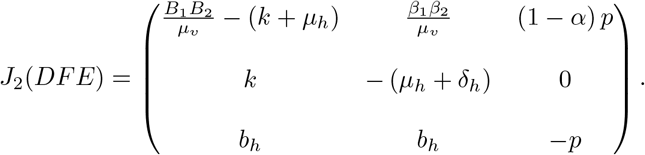

Consider 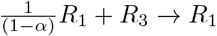, we obtain

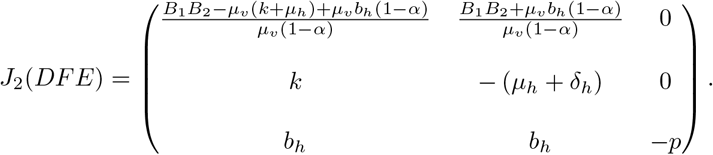

From column 3, the eigenvalue is *ϱ*_4_ = *−p <* 0. The remaining two eigenvalues are given by the 2 *×* 2 matrix

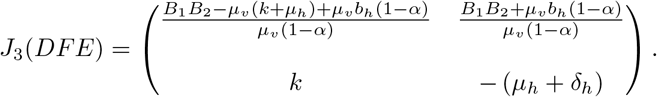

Consider 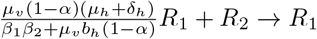, it follows that

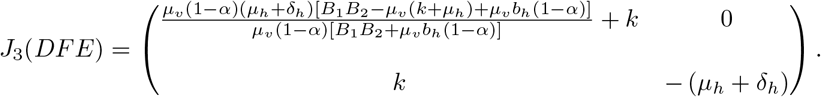

Hence, the eigenvalues of the matrix *J*_3_(*DFE*) are obtain to be

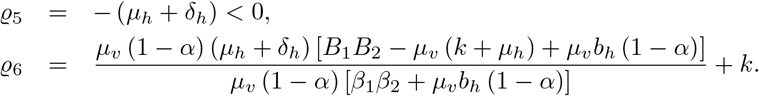

Further simplification of *ϱ*_6_ gives

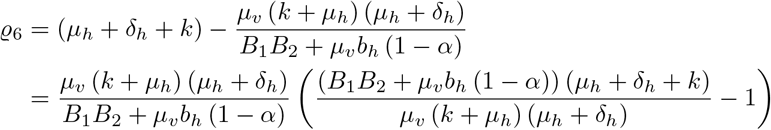

Thus, *ϱ*_6_ *<* 0 if and only if 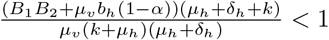. Hence, the disease-free state of the model system is locally asymptotically stable when the above condition is satisfied.

Observe that for *α* = 1, we have

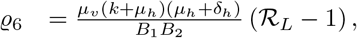

leading to the following result.

### Theorem 1.

*For α* = 1, *the disease-free equilibrium of the system is locally asymptotically stable if ℛ*_*L*_ *<* 1 *and unstable if ℛ*_*L*_ *>* 1.

Next, we will apply the approach of Castillo-Chavez et al [22] to prove the global stability of the disease-free equilibrium. The approach is defined in the theorem below.

### Theorem 2.

*If a model system can be written in the form:*

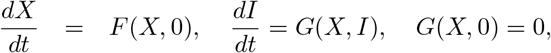

*where X ∈ ℜ*^*m*^ *denotes the number of uninfected compartments and I ∈ ℜ*^*n*^, *denotes the number of infected compartments, including latent, exposed, and acute individuals. U* (*X*^*⋆*^, 0) *denotes the disease-free equilibrium of the system. Then the conditions* (𝔥_1_) *and* (𝔥_2_) *must be satisfied to guarantee local asymptotic stability*.

𝔥_1_ : *For* 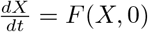, *X*^*⋆*^ *is globally asymptotically stable*.

𝔥_2_ : *G*(*X, I*) = *AI − Ĝ*(*X*, 0) *≥* 0 *for* (*X, I*) *∈* Δ, *where A* = *D*_*i*_*G*(*X*^*⋆*^, 0) *is a Metzler matrix (the off-diagonal elements of A are non-negative) and* Δ *is the region where the model makes biological sense and mathematically well-posed. Then the fixed point U*_0_ = (*X*^*⋆*^, 0) *is globally asymptotically stable equilibrium of the Chagas infection model provided ℛ*_0_ *<* 1.

### Theorem 3.

*The disease-free equilibrium*

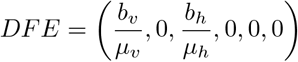

*is globally asymptotically stable if the conditions* (𝔥_1_) *and* (𝔥_2_) *are satisfied. Proof*. From the model system, we have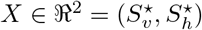 and 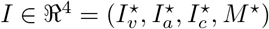. Hence, for condition (𝔥_1_), we have

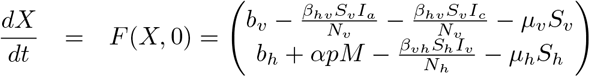

and

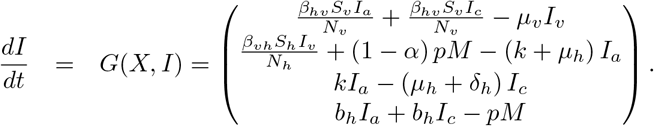

It follows that

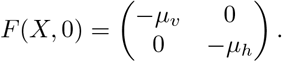

The eigenvaules from the matrix *F* (*X*, 0) are obtained to be

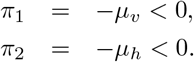

Since all the eigenvalues of the matrix *F* (*X*, 0) are negative, it follows that *X*^*⋆*^ is always globally asymptotically stable. Also, applying Theorem (2) to the Chagas disease model system gives

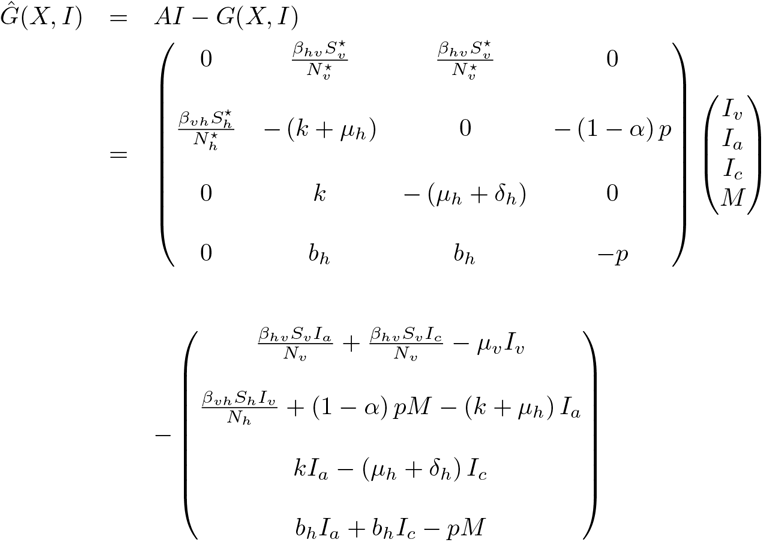

Hence,

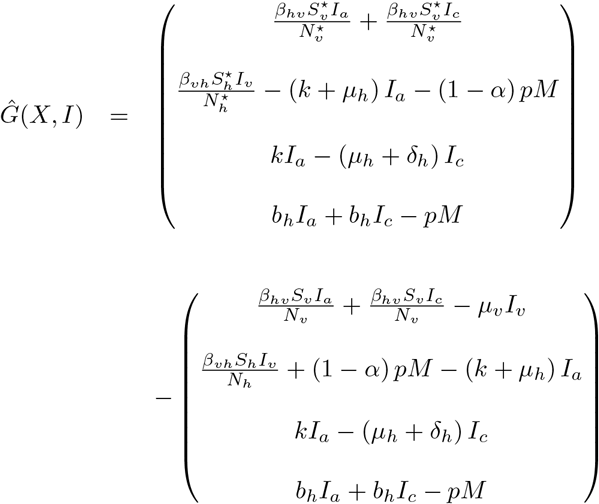

Therefore,

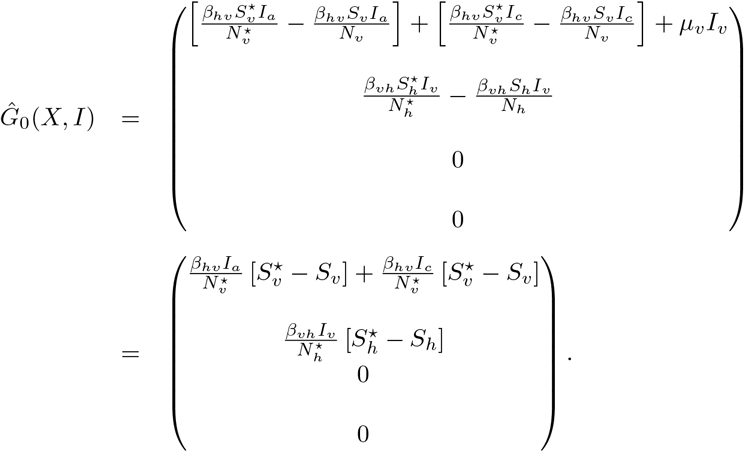

So, *A* is a Metzler matrix with non-negative off-diagonal elements. We observed that

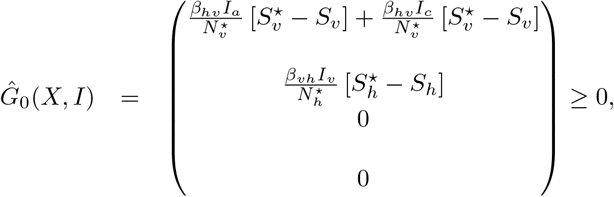

because 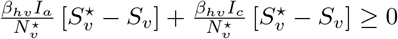 and 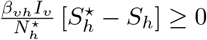. Therefore, the disease-free equilibrium *DFE* is globally asymptotically stable.

## Numerical Simulation Results

In this section, we show the analysis of the numerical simulation of the proposed model. We used literature values and assumed some parameter values to conduct numerical simulations using Matlab for the spread of the Chagas disease. The initial conditions of the state variables are given to be *S*_*h*_(0) = 5000, *I*_*a*_(0) = 1000, *I*_*c*_(0) = 4000, *S*_*v*_(0) = 500000, *I*_*c*_(0) = 100000, *M* (0) = 0 and the rest of the parameters and their values are presented in Table 2.

**Table 2.** Parameters of the disease model and their sources.

| Parameter | Value | Source |
| --- | --- | --- |
| $\beta_{vh}$ | 0.0000032 – 0.0000096 per day | [28] |
| $\beta_{hv}$ | 0.0000012–0.0000036 per day | [28] |
| $\mu_v$ | 0.005 per day | [29] |
| $\mu_h$ | 0.000042 per day | [29] |
| $\delta_h$ | 0.00013– 0.00018 per day | [29] |
| $b_h$ | 70/365 per day | [35] |
| $b_v$ | 183.68–551.04 per day | [34] |
| $k$ | 0.02675 | [29] |
| $p$ | 0.995 | Assumed |
| $r$ | 0.000274 per day | Assumed |
| $\omega$ | 90 - 90% | [20] |

### Simulation and Scenario Analysis

In this section, we used a mathematical model to conduct numerical simulations that depict the dynamics of Chagas disease. These simulations encompassed various scenarios, allowing us to observe how different conditions influence the congenital transmission of Chagas disease. This, in turn, provided valuable insights for optimizing control strategies for Chagas disease. Past approaches for managing the disease have included vector control and early treatment of newborns born to infected mothers. Consequently, we explored scenarios with different newborn treatment rates (*r*), treatment efficacy (*ω*), and varying transmission rates from vectors to humans (*β*_vh_) and humans to vectors (*β*_hv_). All other parameters and values are given in Table 2.

First, we examined how varying *α*, which is the product of the newborn treatment rate (*r*) and the treatment efficacy (*ω*), affects the infected population. Essentially, *α* represents the rate at which newborns born to infected mothers transition from their initial infected state to the susceptible healthy class after receiving proper treatment. Initially, we set *α* to 0.4125, a value determined through a systematic analysis of the reproduction number (*ℛ*_*c*_). We plotted *ℛ*_*c*_ against different combinations of newborn treatment rates (*r*) and treatment efficacy (*ω*) values. Our objective was to identify parameter combinations that resulted in a realistic range of *ℛ*_*c*_ values while ensuring *ℛ*_*c*_ remained below 1. This specific baseline value (*α* = 0.4125) was achieved with newborn treatment rate (*r*) and treatment efficacy (*ω*) values of 0.75 and 0.55, respectively. We also modified *α* by increasing and decreasing it in increments of 25%, 50%, and 75% to examine how varying *α* values affected the spread and incidence of the disease. This led to seven different *α* values: the baseline *α* of 0.4125, a 25% increase (*α* = 0.5156), a 25% decrease (*α* = 0.3094), a 50% increase (*α* = 0.6188), a 50% decrease (*α* = 0.2063), a 75% increase (*α* = 0.7219), and a 75% decrease (*α* = 0.1031). Figure 2 illustrates the population of infected individuals over a 15-year period given these *α* values. The graph exhibits an exponential growth pattern from left to right, which was consistent across all variations of *α* values, signifying an increase in the acutely infected

**Fig 2.**
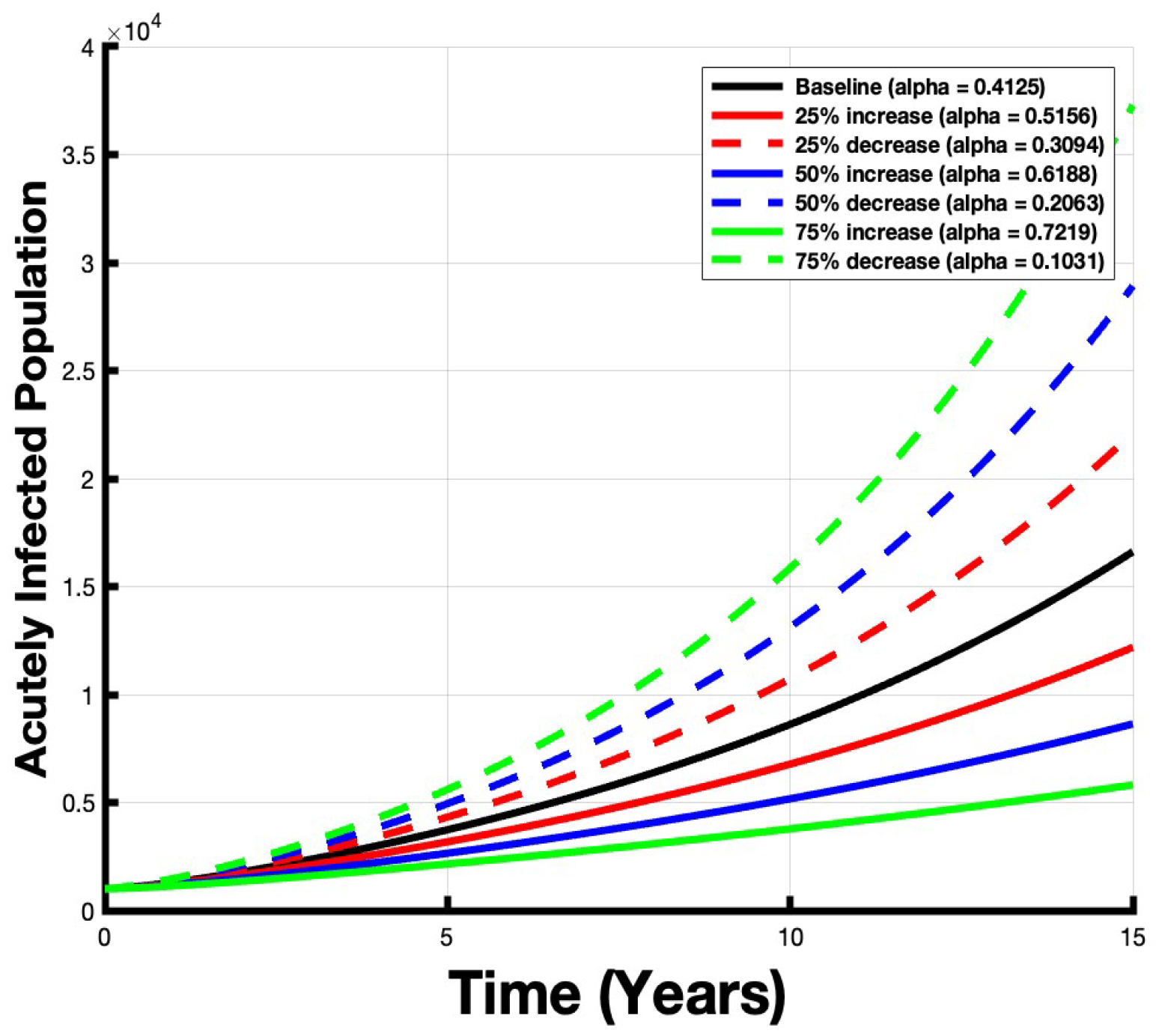
The acutely infected population increases over a period of 15 years for all values of *α*. Relative to the baseline, increasing *α* leads to a decrease in infections, while decreasing *α* corresponds to an increase in infections.

population in each scenario over 15 years. We calculated the area under the curve (AUC), providing a measure of the infected population over time for each *α* value.

The respective AUC values for the different scenarios were as follows:

- Baseline scenario (*α* = 0.4125): AUC = 102,232
- 25% increase in *α* (*α* = 0.5156): AUC = 80,519
- 25% decrease in *α* (*α* = 0.3094): AUC = 127,568
- 50% increase in *α* (*α* = 0.6188): AUC = 61,928
- 50% decrease in *α* (*α* = 0.2063): AUC = 157,102
- 75% increase in *α* (*α* = 0.7219): AUC = 46,023
- 75% decrease in *α* (*α* = 0.1031): AUC = 191,497

The calculated percentage changes represent the difference in the infected population compared to the baseline scenario, illustrating the impact of adjusting *α* values on disease dynamics. The percentage change values were as follows:

- Percentage change for a 25% increase in *α*: *−*21.2%
- Percentage change for a 25% decrease in *α*: 24.7%
- Percentage change for a 50% increase in *α*: *−*39.4%
- Percentage change for a 50% decrease in *α*: 53.6%
- Percentage change for a 75% increase in *α*: *−*54.9%
- Percentage change for a 75% decrease in *α*: 87.3%

Interpreting these results, a 25% increase in *α* correlated with a 21.2% decrease in the infected population relative to the baseline. Conversely, a 25% decrease in *α* corresponded to a 24.7% increase in the infected population. Similarly, a 50% increase in *α* resulted in a 39.4% reduction in the infected population, signifying significant progress in disease management. Meanwhile, a 50% decrease in *α* led to a 53.6% increase in the infected population. A 75% increase in *α* correlated with a 54.9% reduction in the infected population, while a 75% decrease in *α* resulted in an 87.3% increase in infections. These results underscore how *α*, representing newborn treatment rate and treatment effectiveness, influences disease spread. Ultimately, increasing *α* leads to a decrease in the infected population, while decreasing *α* leads to an increase in the infected population.

### Further Exploring Impact of *α*

To further explore the impact of *α*, we considered scenarios by setting *α* to its minimum (*α* = 0) and maximum (*α* = 1) values. An *α* value of 0 would mean that none of the newborns from infected mothers would transition from their infected state to a healthy state. This could be explained by two situations: the newborns from infected mothers would either not be getting treated (*r* = 0), or they would receive treatment that did not work (*ω* = 0). On the other hand, an *α* value of 1 would mean that all of the newborns from infected mothers would transition from their infected state to a healthy state, indicating that all of the newborns received treatment (*r* = 1) which was 100% effective (*ω* = 1). Figure 3 shows the population of infected individuals over a period of 15 years when *α* is at its minimum value of 0. This AUC for this case is 231,509 individuals; this corresponds to a 126.3% increase in the infected population. In contrast, Figure 4 shows the infected population’s trajectory over 15 years at the maximum *α* value of 1. The AUC for *α* = 1 is 13,735 individuals. Comparing this with the earlier baseline scenario, the infected population’s percentage change stands at *−*86.5%. This considerable reduction from the baseline suggests that maximizing the newborn treatment rate and efficacy rate is effective in reducing the burden of Chagas disease.

**Fig 3.**
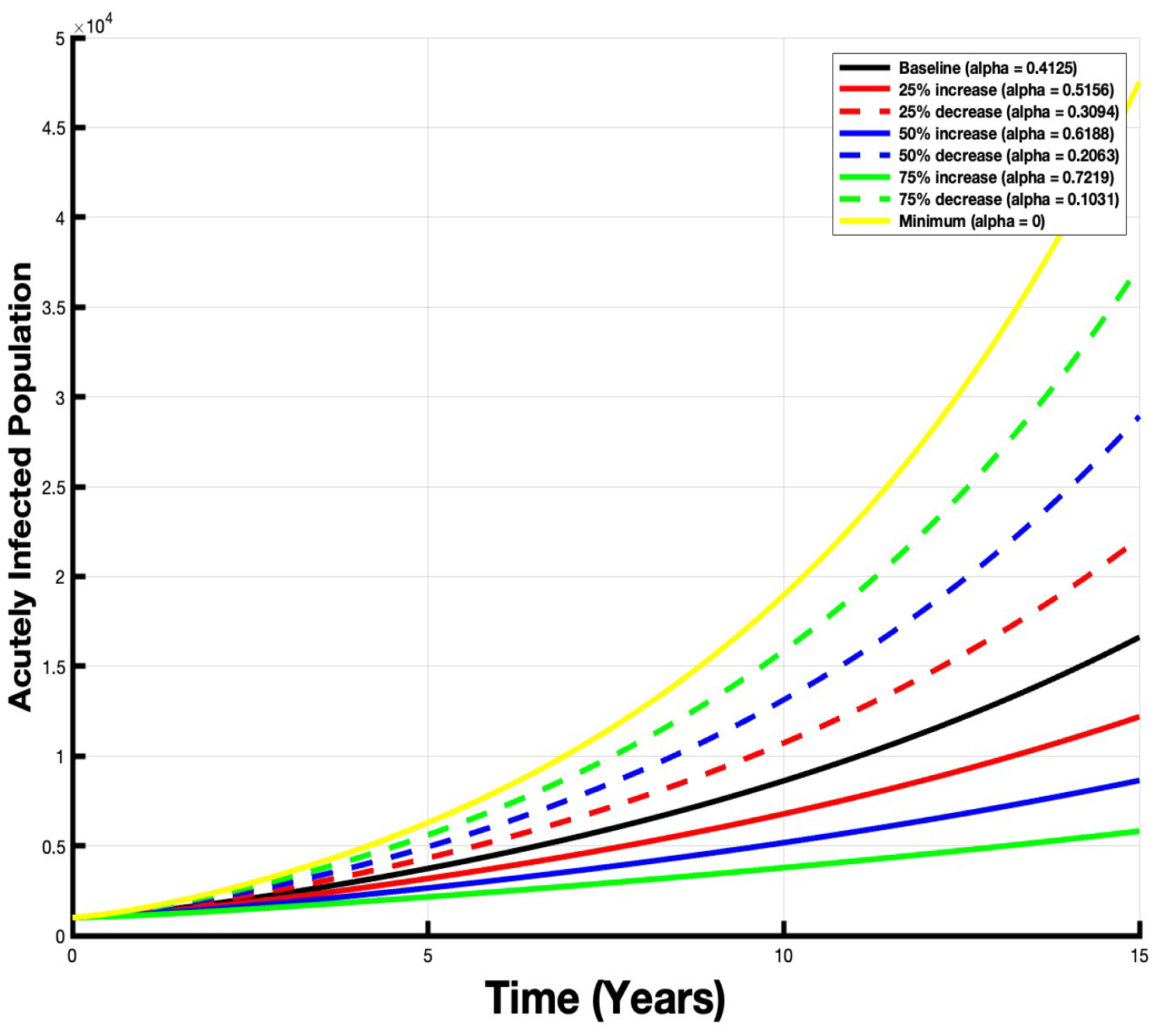
The number of acutely infected individuals experiences its highest increase over 15 years when the *α* value is set to 0.

**Fig 4.**
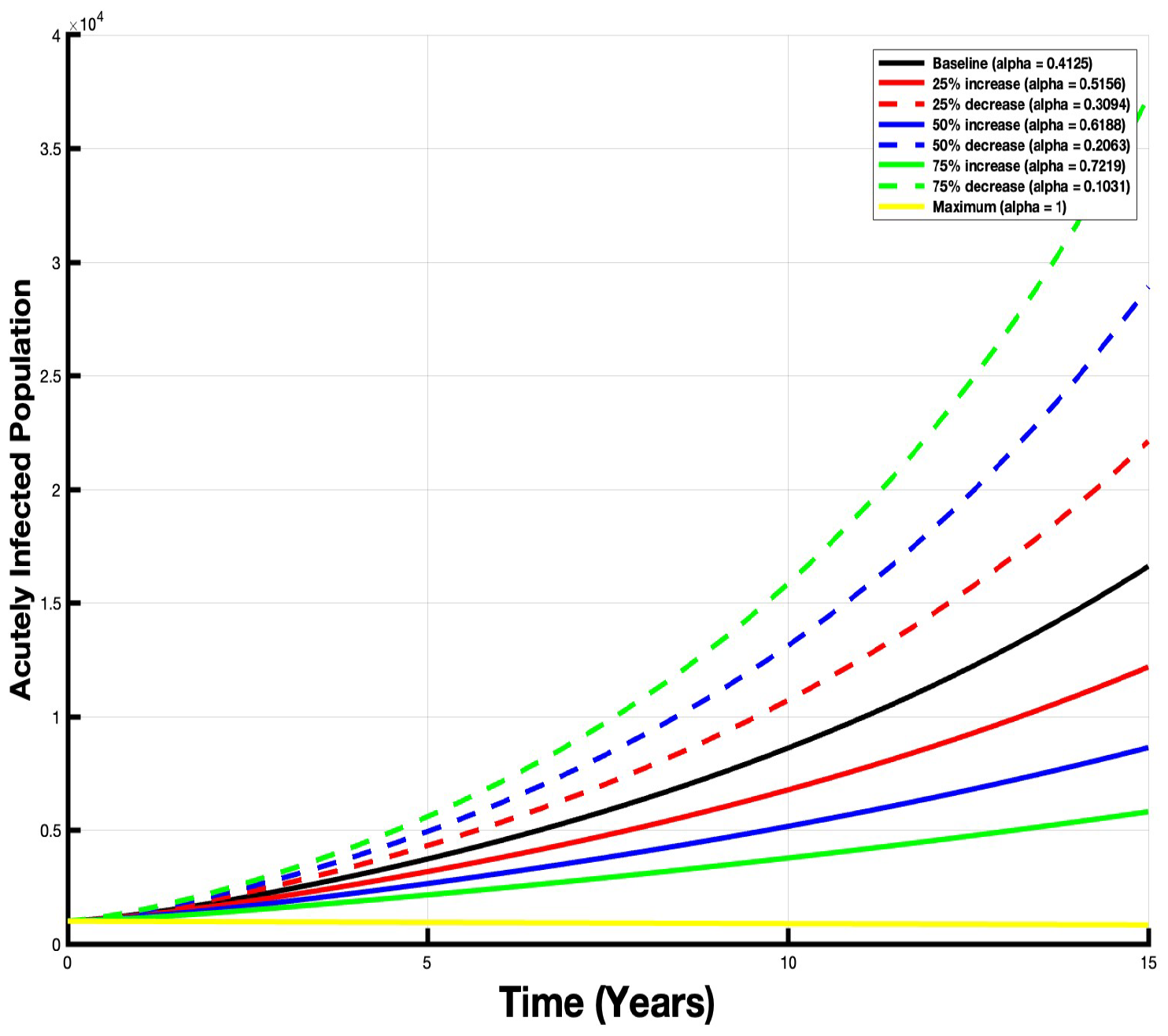
The number of acutely infected individuals decreases over 15 years when *α* = 1.

### Impact of Vector Control

Finally, the influence of vector control on Chagas disease dynamics is considered. In Chagas disease transmission dynamics, two parameters describe the interactions between vectors and humans: *β*_*υh*_, representing the transmission rate from infected vectors to susceptible humans, and *β*_*hυ*_, signifying the transmission rate from infected humans to susceptible vectors. The baseline values for these parameters are given in Table 2.

To investigate the dynamics of Chagas disease when there is no transmission of the disease from vectors to humans, the impact of varying *β*_*υh*_ on the acutely infected population is explored. Figure 5 illustrates the dynamics of the acutely infected population under the same baseline *α* value (*α* = 0.4125) but with different values for *β*_*υh*_. In the baseline scenario (*α* = 0.4125 and *β*_*υh*_ = 0.0000036), the infected population remains considerable, as indicated by the area under the curve of 102,231.8543.

**Fig 5.**
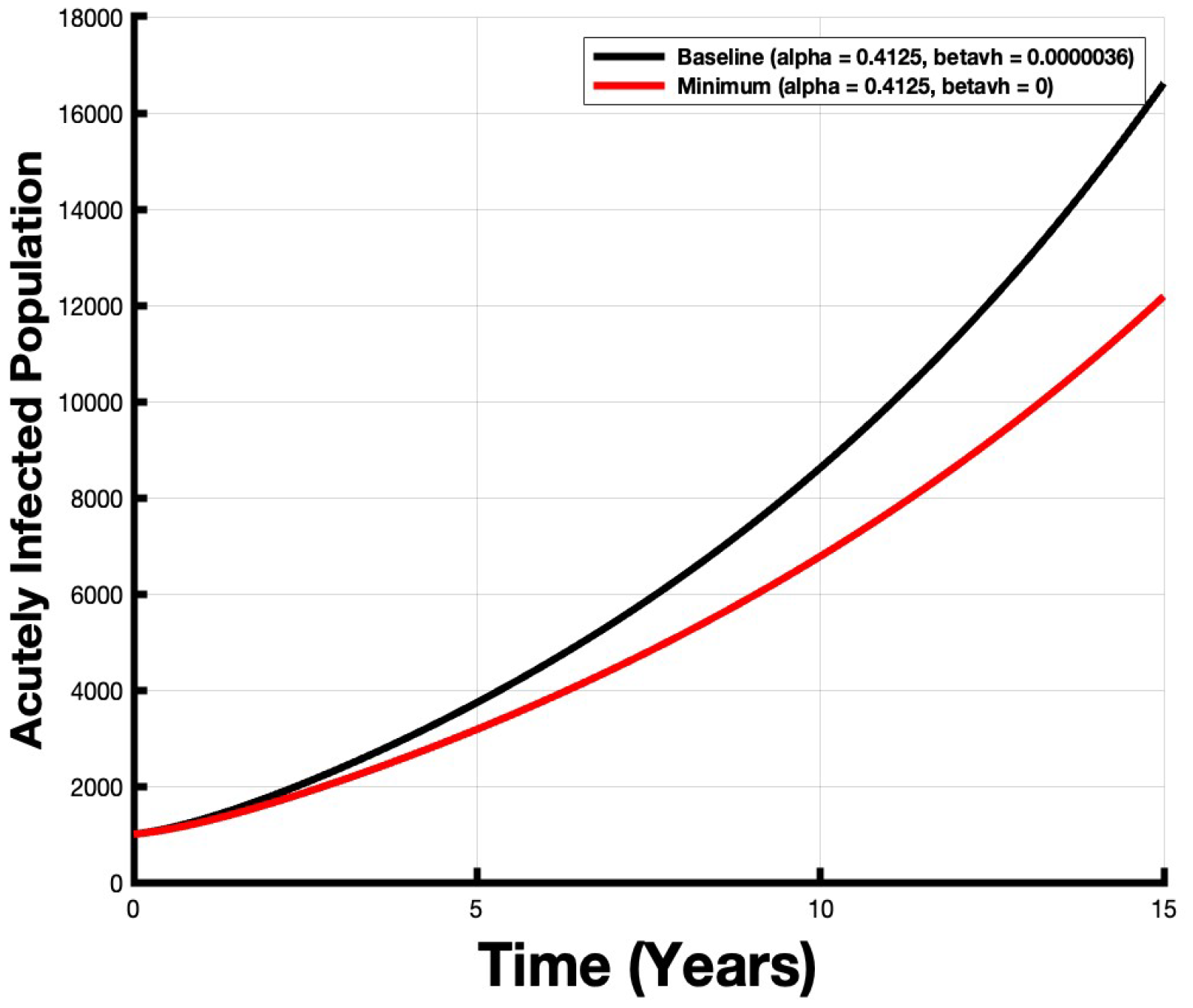
The number of acutely infected individuals decreases over 15 years when *β*_*υh*_ = 0.

However, in the scenario where *β*_*υh*_ is set to zero, the infected population is significantly reduced, reflected by the much lower area under the curve of 80,519. This substantial 21% decrease in the infected population emphasizes the importance of vector control measures in reducing the spread and impact of Chagas disease.

## Discussion

The results from our mathematical model that investigates the dynamics of Chagas disease provide valuable information about the factors influencing the congenital transmission of this disease. In this section, we discuss the implications of our findings and their significance for strengthening control strategies for Chagas disease.

The newborn treatment rate (*r*) and treatment efficacy (*ω*) are represented by *α*, which plays a significant role in shaping the dynamics of Chagas disease. Our results highlight that changing *α* has a significant impact on the spread of the disease. Increasing *α* results in a reduction in the infected population by 21.2%, 39.4%, and 54.9% for 25%, 50%, and 75% *α* increases, respectively. Conversely, decreasing *α* by the same percentages leads to an increase in the infected population by 24.7%, 53.6%, and 87.3%. This finding highlights the importance of effective treatment of newborns born to infected mothers as it can greatly reduce the burden of Chagas disease. Additionally, the results show the impact of minimizing (*α* = 0) and maximizing newborn treatment rates and efficacy (*α* = 1). Minimizing *α*, which suggests that newborns from infected mothers would either not receive treatment (*r* = 0) or receive treatment that does not work (*ω* = 0), led to a 126.4% increase in the infected population. On the other hand, maximizing *α* led to an 86.5% reduction in the infected population. When *α* = 1, this guarantees the maximum impact and effectiveness, as it implies that 100% of newborns are receiving treatment, and the treatment is 100% effective. However, it’s essential to acknowledge that such a scenario is realistic but not sufficient to eradicate Chagas disease, as 13.4% of infections continue to exist despite the most optimal treatment scenario. Thus, finding a balance between treatment rates and efficacy that is both effective and achievable is crucial in disease management.

While our results underscore the significant impact of treatment rate (*r*) and treatment efficacy (*ω*) in influencing the spread of Chagas disease, it is crucial to recognize that these factors alone are necessary but not sufficient for eliminating the disease burden. A more comprehensive approach that includes newborn therapy and vector control strategies is crucial to effectively combat Chagas disease. This is important because Chagas disease primarily spreads through the triatomine bugs, which serve as vectors for the Trypanosoma cruzi parasite. These vectors play a pivotal role in disease transmission, and their control is essential for reducing human infections. Our simulations demonstrate the substantial impact of reducing the transmission of the disease from vectors to humans by setting *β*_*υh*_ to zero. The adjustment, setting *β*_*υh*_ to zero while maintaining the baseline *α* value (*α* = 0.4125), resulted in a significant 21% decrease in the infected population compared to the scenario where *β*_*υh*_ = 0.0000036. This underscores how essential vector control measures are. Vector control includes various strategies, such as insecticide spraying, addressing poor housing conditions, and initiating educational programs to reduce human-vector contact. Implementing vector control strategies can effectively complement the efforts to improve treatment rates and efficacy and further reduce disease transmission.

Based on our results, it is clear that a multifaceted approach is imperative for managing Chagas disease. This approach includes increasing the newborn treatment, enhancing the treatment efficiency, and implementing vector control measures.

High treatment rates and treatment efficacy are important for controlling and reducing the burden of Chagas disease; however, they do not address vector-borne transmission. Hence, it becomes clear that vector control has to be part of the multifaceted approach to mitigate Chagas disease transmission. Public health interventions should consider these varied methods of disease control to develop exhaustive strategies that regulate both congenital transmission and vector-to-human transmission routes.

Further research in Chagas disease control should focus on developing integrated approaches that include both treatment and vector control strategies. Based on our research, we recommend initiatives that raise awareness about Chagas disease and promote early diagnosis and treatment. With these combined efforts, the burden of Chagas disease can be significantly reduced, protecting many people from its harmful effects.

## Data Availability

All data produced in the present work are contained in the manuscript

## Notes

### Competing Interest Statement

The authors have declared no competing interest.

### Funding Statement

This study did not receive any funding

## References

1. Benatar AF, Danesi E, Besuschio SA, Bortolotti S, Cafferata ML, Ramirez JC, Albizu CL, Scollo K, Baleani M, Lara L, Agolti G, Seu S, Adamo E, Lucero RH, Irazu L, Rodriguez M, Poeylaut-Palena A, Longhi SA, Esteva M, Althabe F, Rojkin F, Bua J, Sosa-Estani S, Schijman AG; Congenital Chagas Disease Study Group. Prospective multicenter evaluation of real-time PCR Kit prototype for early diagnosis of congenital Chagas disease. EBioMedicine. 2021 Jul;69:103450. doi: 10.1016/j.ebiom.2021.103450. Epub 2021 Jun 26. PMID: 34186488; PMCID: PMC8243352.

2. Bern C, Martin DL, Gilman RH. Acute and congenital Chagas disease. Adv Parasitol. 2011;75:19–47. doi: 10.1016/B978-0-12-385863-4.00002-2. PMID: 21820550.

3. Bern C, Montgomery SP, Herwaldt BL, et al. Evaluation and Treatment of Chagas Disease in the United States: A Systematic Review. JAMA. 2007;298(18):2171–2181. doi:10.1001/jama.298.18.2171

4. Bonney K. M. (2014). Chagas disease in the 21st century: a public health success or an emerging threat?. Parasite (Paris, France), 21, 11. 10.1051/parasite/2014012

5. Bustos PL, Milduberger N, Volta BJ, Perrone AE, Laucella SA and Bua J (2019) Trypanosoma cruzi Infection at the Maternal-Fetal Interface: Implications of Parasite Load in the Congenital Transmission and Challenges in the Diagnosis of Infected Newborns. Front. Microbiol. 10:1250. doi: 10.3389/fmicb.2019.01250

6. Carlier Y, Truyens C. Congenital Chagas disease as an ecological model of interactions between Trypanosoma cruzi parasites, pregnant women, placenta and fetuses. Acta Trop. 2015 Nov;151:103–15. doi: 10.1016/j.actatropica.2015.07.016. Epub 2015 Aug 17. PMID: 26293886.

7. Cevallos AM, Hernández R. Chagas’ disease: pregnancy and congenital transmission. Biomed Res Int. 2014;2014:401864. doi: 10.1155/2014/401864. Epub 2014 May 15. PMID: 24949443; PMCID: PMC4052072.

8. da Nóbrega, A. A., de Araújo, W. N., & Vasconcelos, A. (2014). Mortality due to Chagas disease in Brazil according to a specific cause. The American journal of tropical medicine and hygiene, 91(3), 528–533. 10.4269/ajtmh.13-0574

9. Fabbro DL, Danesi E, Olivera V, Codebó MO, Denner S, Heredia C, Streiger M, Sosa-Estani S. Trypanocide treatment of women infected with Trypanosoma cruzi and its effect on preventing congenital Chagas. PLoS Negl Trop Dis. 2014 Nov 20;8(11):e3312. doi: 10.1371/journal.pntd.0003312. PMID: 25411847; PMCID: PMC4239005.

10. García-Huertas P, Cardona-Castro N. Advances in the treatment of Chagas disease: Promising new drugs, plants and targets. Biomed Pharmacother. 2021 Oct;142:112020. doi: 10.1016/j.biopha.2021.112020. Epub 2021 Aug 12. PMID: 34392087.

11. Hector Freilij, & Jaime Altcheh. (1995). Congenital Chagas’ Disease: Diagnostic and Clinical Aspects. Clinical Infectious Diseases, 21(3), 551–555. http://www.jstor.org/stable/4458866

12. Jurado Medina L, Chassaing E, Ballering G, Gonzalez N, Marqúe L, Liehl P, Pottel H, de Boer J, Chatelain E, Zrein M, Altcheh J. Prediction of parasitological cure in children infected with Trypanosoma cruzi using a novel multiplex serological approach: an observational, retrospective cohort study. Lancet Infect Dis. 2021 Aug;21(8):1141–1150. doi: 10.1016/S1473-3099(20)30729-5. Epub 2021 Apr 6. PMID: 33836157.

13. Klotz, S. A., Dorn, P. L., Mosbacher, M., & Schmidt, J. O. (2014). Kissing bugs in the United States: risk for vector-borne disease in humans. Environmental health insights, 8(Suppl 2), 49–59. 10.4137/EHI.S16003

14. Lidani KCF, Andrade FA, Bavia L, Damasceno FS, Beltrame MH, Messias-Reason IJ and Sandri TL (2019) Chagas Disease: From Discovery to a Worldwide Health Problem. Front. Public Health 7:166. doi: 10.3389/fpubh.2019.00166

15. Llenas-García J, Wikman-Jorgensen P, Gil-Anguita C, Ramos-Sesma V, Torrús-Tendero D, Martínez-Goñi R, Romero-Nieto M, García-Abellán J, Esteban-Giner MJ, Antelo K, Navarro-Cots M, Buñuel F, Amador C, García-García J, Gascón I, Telenti G, Fuentes-Campos E, Torres I, Gimeno-Gascón A, RúIz-García MM, Navarro M, Ramos-Rincón JM. Chagas disease screening in pregnant Latin American women: Adherence to a systematic screening protocol in a non-endemic country. PLoS Negl Trop Dis. 2021 Mar 24;15(3):e0009281. doi: 10.1371/journal.pntd.0009281. PMID: 33760816; PMCID: PMC8021187.

16. Meymandi, S., Hernandez, S., Park, S. et al. Treatment of Chagas Disease in the United States. Curr Treat Options Infect Dis 10, 373–388 (2018). 10.1007/s40506-018-0170-z

17. Nguyen T, Waseem M. Chagas Disease. [Updated 2021 Nov 7]. In: StatPearls [Internet]. Treasure Island (FL): StatPearls Publishing; 2022 Jan-. Available from: https://www.ncbi.nlm.nih.gov/books/NBK459272/

18. Picado, A., Cruz, I., Redard-Jacot, M., Schijman, A. G., Torrico, F., Sosa-Estani, S., Katz, Z., & Ndung’u, J. M. (2018). The burden of congenital Chagas disease and implementation of molecular diagnostic tools in Latin America. BMJ global health, 3(5), e001069. 10.1136/bmjgh-2018-001069

19. Requena-Méndez A, Albajar-Viñas P, Angheben A, Chiodini P, Gascón J, Muñoz J; Chagas Disease COHEMI Working Group. Health policies to control Chagas disease transmission in European countries. PLoS Negl Trop Dis. 2014 Oct 30;8(10):e3245. doi: 10.1371/journal.pntd.0003245. PMID: 25357193; PMCID: PMC4214631.

20. Soriano-Arandes A, Angheben A, Serre-Delcor N, Treviño-Maruri B, Gómez I Prat J, Jackson Y. Control and management of congenital Chagas disease in Europe and other non-endemic countries: current policies and practices. Trop Med Int Health. 2016 May;21(5):590–6. doi: 10.1111/tmi.12687. Epub 2016 Mar 17. PMID: 26932338.

21. Steverding, D. The history of Chagas disease. Parasites Vectors 7, 317 (2014). 10.1186/1756-3305-7-317

22. Castillo-Chavez, C., Blower, S., Van den Driessche, P., Kirschner, D. and Yakubu, A. Mathematical approaches for emerging and reemerging infectious diseases: an introduction. Springer Science and Business Media, (2002)

23. Oduro B., Grijalva M.J., Just W. (2018). Models of disease vector control: When can aggressive initial intervention lower long-term cost? Bulletin of Mathematical Biology, 80 (4), pp. 788–824

24. Oduro B., Grijalva M.J., Just W. (2019). A model of insect control with imperfect treatment. Journal of Biological Dynamics, 13 (NO. 1), pp. 518-537, 10.1080/17513758.2019.1640293

25. Spagnuolo, A. M., Shillor, M., Stryker, G. A. (2011). A model for Chagas disease with controlled spraying. Journal of Biological Dynamics 5 299–317.

26. Raimundo, S. M., Massad, E., and Yang, H. M. (2010). Modelling congenital transmission of Chagas’ disease. Biosystems, 99(3), 215–222. 10.1016/j.biosystems.2009.11.005

27. Coffield DJ Jr, Spagnuolo AM, Shillor M, Mema E, Pell B, Pruzinsky A, et al. (2013) A Model for Chagas Disease with Oral and Congenital Transmission. PLoS ONE 8(6): e67267. 10.1371/journal.pone.0067267

28. Oduro B., Naandam S., Mock D., Miloua A., Chataa P. (202?)The impact of continuous insecticide spraying on the spread of Chagas disease. Applied Mathematical Analysis and Computation: Springer Proceedings in Mathematics & Statistics.

29. Cruz-Pachecoa, G., Estevab L., Vargas, C. (2012). Control measures for Chagas disease. Mathematical Biosciences 237 49–60.

30. Diekmann, O., Heesterbeek, J. A. P, Britton, T. (2012). Mathematical Tools for Understanding Infectious Disease Dynamics. Kindle Edition. Princeton University Press.

31. Driessche, P. V., Watmough, J. (2002). Reproduction numbers and sub-threshold endemic equilibria for compartmental models of disease transmission. Mathematical Biosciences 180 29–48.

32. Heesterbeek, J. A. P. (2002). A brief history of R0 and a recipe for its calculation. Acta Biotheo 50 189—204.

33. Korobeinikov, A. (2007). Global properties of infectious disease models with nonlinear incidence. Bull Math Biol 69 1871–1886.

34. Spagnuolo, A. M., Shillor, M., Kingsland, L., Thatcher, A., Toeniskoetter, M., Wood, B. (2012). A logistic delay differential equation model for Chagas disease with interrupted spraying schedules. Journal of biological dynamics 6(2) 377–394.

35. Hidayat, D., Nugraha, E. S., Nuraini, N. (2018). A mathematical model of Chagas disease transmission. In AIP Conference Proceedings 1937(1) 020008. AIP Publishing LLC.

